# Cerebrospinal fluid levels of NfM in relation to NfL and pNfH as prognostic markers in amyotrophic lateral sclerosis

**DOI:** 10.1101/2024.07.09.24310135

**Authors:** Jennie Olofsson, Sofia Bergström, Sára Mravinacová, Ulf Kläppe, Linn Öijerstedt, Henrik Zetterberg, Kaj Blennow, Caroline Ingre, Peter Nilsson, Anna Månberg

**Author notes:** Correspondence to: Dr. Anna Månberg Dept of Protein Science, KTH Royal Institute of Technology Tomtebodavägen 23, 171 65 Solna, Sweden.

## Abstract

**Objective:** To evaluate the prognostic potential of neurofilament medium chain (NfM) in CSF from patients with ALS and explore its relationship with the extensively studied neurofilament light chain (NfL) and phosphorylated heavy chain (pNfH).

**Method:** CSF levels of NfL, NfM, and pNfH were analysed in 235 samples from patients with ALS, ALS mimics, and healthy controls in a well-characterized cohort from Karolinska ALS Clinical Research Centre in Stockholm, Sweden. NfM levels were analysed using an antibody-based suspension bead-array and NfL and pNfH levels were measured using standard ELISA. Clinical data, including ALS Revised Functional Rating Scale (ALSFRS-R), and survival outcomes were utilized for disease progression estimations.

**Result:** Increased NfM levels were observed in patients with ALS compared with mimics and healthy controls, consistent with the results observed for NfL and pNfH. Similar to NfL and pNfH, significant associations of NfM levels with disease progression were found, with higher levels in fast progressors compared with slow progressors, both in total progression and the ALSFRS-R subscores. For all neurofilaments, higher survival probability was observed for patients with low CSF levels. Additionally, all three proteins showed similar predictive performance for disease progression rates (AUC 0.85-0.86). Although there was no statistical difference between the curves, combining them slightly improved the performance (AUC 0.86-0.87).

**Conclusion:** Based on this cross-sectional study, the prognostic value provided by NfM aligns with the more established markers, NfL and pNfH. However, additional investigations with independent cohorts and longitudinal studies are needed to further explore the potential added value of NfM.

## Introduction

Amyotrophic lateral sclerosis (ALS) is a fatal neurodegenerative disease that specifically affects upper and lower motor neurons. The progression of the disease is often rapid, and individuals diagnosed with ALS typically survive two to three years after symptom onset (1). However, the disease progression rate can vary and a small subset of patients surpass the ten-year survival mark after onset of the disease (2, 3).

Numerous studies have consistently reported increased neurofilament light chain (NfL) and phosphorylated neurofilament heavy chain (pNfH) levels in both cerebrospinal fluid (CSF) and blood from patients with ALS when compared with healthy controls and disease mimics, such as polyneuropathy, myopathy, myositis, hereditary spastic paraplegia, benign fasciculations and other diseases, where initial symptoms mimicked those of ALS (4–10). Many of these studies have found associations between increased neurofilament levels and disease progression rates, as well as survival outcomes. Furthermore, correlations between NfL and pNfH levels and the revised ALS Functional Rating Scale (ALSFRS-R) score have been identified (6, 11–13). Notably, NfL and pNfH have been utilized as secondary endpoints in clinical trials (14, 15).

While many studies have examined NfL and pNfH levels in ALS, research investigating the protein levels of neurofilament medium chain (NfM) in relation to the disease is less explored. Increased NfM levels in plasma and CSF among patients with ALS compared with controls, and associations to disease progression, have previously been reported (16–19). Also, increased NfM levels have been found in CSF from patients with frontotemporal dementia (FTD), a disease with overlapping pathological features with ALS (20, 21). However, these studies have not explored the levels of NfM in CSF among patients with ALS and its relationship to disease progression and survival, nor the associations between the different neurofilament proteins.

In this study, our primary objective was to evaluate the prognostic potential of CSF NfM levels within ALS and investigate the relationship with NfL and pNfH. Additionally, we aimed to determine whether the combined use of the different neurofilaments outperforms their individual ability to predict disease progression and survival. Lastly, we aimed to examine the ALSFRS-R subscores in relation to the protein levels separately, to increase the understanding of specific aspects of disease manifestation and progression.

## Method

### Participants and clinical data

Study participants were prospectively recruited (2016–2021) at the Karolinska ALS Clinical Research Centre, a tertiary clinic that cares for all patients with ALS in Stockholm. Patients diagnosed with ALS according to the Gold Coast Criteria, with a spinal or bulbar onset, and with available CSF samples drawn within 90 days of diagnosis, were included in the analysis (n = 182). Of the 182 patients with ALS, 8 patients only had signs of lower motor neuron involvement and thus diagnosed as progressive muscular atrophy (PMA). Additionally, healthy controls (n = 11) and ALS mimics (n = 42) were included in the study. ALS mimics were individuals evaluated at the ALS Clinical Research Centre with suspicion of ALS, but who were later diagnosed with other neurological diseases (such as polyneuropathy, multiple sclerosis, spastic paraparesis, myelopathy, Parkinson’s disease, neuroborreliosis, spinal stenosis, benign fasciculations, etc.) (**Supplementary table 1**). The age and sex distribution per group (ALS, mimics, healthy controls) is presented in **Table 1**.

**Table 1:** Sample demographics.

|  | **ALS** | **Mimics** | **Healthy**<br>**controls** | ***p*-value** |
| --- | --- | --- | --- | --- |
| **Number of individuals** | 182 | 42 | 11 |  |
| **Age [median (IQR)]** | 67 (59-74) | 71 (63-75) | 60 (56-65) | 0.035<sup>a</sup> |
| **Sex [F/M]** | 78/104 | 10/32 | 5/6 | 0.069<sup>b</sup> |

<sup>a</sup>Kruskal Wallis rank sum test. Mann-Whitney U test for pairwise comparison (ALS vs.
healthy controls, $p=0.03$ ; Mimics vs. healthy controls, $p=0.009$ ). <sup>b</sup>Chi-square test. IQR =
interquartile range.

This study was approved by the Swedish Ethical Review Authority (Dnrs 2014/1815-31/4, 2017/1895-31/1, 2009/2107-31/2, 2017/952-31). The study followed the ethical principles of the Declaration of Helsinki and all participants provided written consent.

Functional disability was evaluated through the utilization of the ALSFRS-R (22). ALSFRS-R scores from multiple visits were available for most of the patients (mean number of visits = 8, range 1-28). Average disease progression rate was calculated as the ALSFRS-R score at diagnosis minus ALSFRS-R score at last visit divided by number of months between the two measurements. Scores from at least three occasions were required to assess the average disease progression rate, which were available for 147 (81%) of the patients. Item two of the ALSFRS-R is assessing salivation and was omitted in the analysis due to the potential influence of medications on salivation.

The patients with ALS were stratified into progression groups based on the average disease progression rate. Individuals with an average disease progression rate faster than 1.5 points/month was considered fast progressors, while individuals with an average disease progression rate slower than 0.4 points/month were considered slow progressors. In addition to the total ALSFRS-R scores, we investigated the subscores of the ALSFRS-R domains, i.e., bulbar, fine motor, gross motor, and respiratory. Survival status was noted from date of diagnosis until the date of death or invasive ventilation, whichever came first. Patients who were still alive and with no invasive ventilation were censored from 20^th^ of October 2023. Progression rate, along with survival time information, for individuals with bulbar and spinal onset is presented in **Table 2**.

**Table 2:** Number of indivudials with ALS per onset, disease progression rate, and survival times.

|  |  | **Spinal onset** | **Bulbar onset** |
| --- | --- | --- | --- |
|  |  | (n=118) | (n=64) |
| **Progression [N (%)]** | *Fast* | 21 (18) | 17 (27) |
|  | *Intermediate* | 66 (56) | 24 (38) |
|  | *Slow* | 13 (11) | 6 (9) |
|  | *Incomplete data* | 18 (15) | 17 (27) |
| **Survival [Mean months (range)]** |  | 27 (1-84) | 17 (1-59) |

Note: fast progression: average disease progression rate $> 1.5$ ; slow progression: average disease progression rate $< 0.4$ .

### Sample collection and protein measurements

CSF was obtained through lumbar puncture, and collected in polypropylene (PP) tubes and centrifuged for ten minutes at 400 g at room temperature. All samples were aliquoted within two hours, directly frozen and stored in −80°C.

NfL and pNfH were measured using sandwich enzyme-linked immunosorbent assays (ELISA). Commercially available ELISA kits were used, pNfH from Euroimmun, Lübeck, Germany, and NfL from UmanDiagnostics, Sweden. NfL concentrations ranged from 168 to 62710 pg/ml, and from 61 to 22060 pg/ml for pNfH. The experimental data for NfL and pNfH in a subset of the patients with ALS, healthy controls, and mimics, was analysed in relation to disease progression and survival in a previous study and the results published separately (11).

NfM levels were measured with the antibody-based suspension bead array technology (23, 24), using a polyclonal rabbit IgG antibody (HPA022485) generated within the Human Protein Atlas project (HPA, www.proteinatlas.org). More details on experimental procedures can be found in a previous publication (24). The anti-NfM antibody has previously been validated using western blots, sandwich assays, and parallel reaction monitoring mass spectrometry (16, 20, 25). The data were normalized to reduce the impact of time delay during data readout. A robust linear regression analysis was conducted (*rlm, MASS*), and the generated residuals were then added to the median signal intensity. This was followed by MA-Individual normalization (26) to reduce potential differences between plates. In the assay measuring NfM levels, the technical coefficient of variation was 2.6% and the inter-assay correlation was ρ=0.98.

### Data analysis and visualizations

All data analysis and visualisations were performed using the open-source software R (version 4.3.2) (27). The protein levels were log2-transformed and standardized via mean centering and unit variance scaling prior to analysis (z-scores).

Kruskal-Wallis rank sum test (*kruskal.test, stats*) was used to test differences across multiple groups. For comparisons between two groups, the Mann-Whitney U test (*wilcox.test, stats*) was utilized. Spearman’s rank correlation (*cor, stats*) was applied to calculate correlations between continuous variables. Pearson’s Chi-squared test (*chisq.test, stats*) was used to investigate differences in sex distribution between the diagnostic groups. P-values < 0.05 were considered statistically significant.

To assess if the protein levels could predict progression rate, we first constructed generalized linear models (*glm, stat*) that considered protein levels as predictors, separately or in combination, and the disease progression rate as the response variable. This was followed by generation of predictive estimates based on the fitted model. Performance was evaluated with Area Under the Curve (AUC) and Receiver Operating Characteristics (ROC) curves (*roc, pROC*), and difference between AUC was assessed using the DeLong test (*roc.test, pROC*).

Kaplan-Meier survival analysis was applied to assess the survival probability based on the levels of the three proteins. In this analysis, the patients were stratified into high, intermediate, and low protein level groups based on terciles, before differences between the high and low level groups were assessed using log-rank test and Kaplan-Meier curves (*survfit, survminer*). Initially, cox-proportional hazard models (*coxph, survival*) were employed to evaluate the risk associated with each cofactor (sex, age, site of onset). However, these models could not be completed due to a violation of the proportional hazards assumption.

## Results

### CSF levels of NfM are higher in patients with ALS compared with controls

Significantly increased NfM levels were observed in patients with ALS compared with healthy controls and with mimics, in line with the results for NfL and pNfH (**Fig. 1A**). Correlations between levels of the three subunits among patients with ALS were strong (NfL-NfM, ρ=0.84; pNfH-NfM, ρ=0.82; pNfH-NfL, ρ=0.86), although slight differences in assay sensitivity were observed (**Fig. 1B**). No significant differences between patients with spinal and bulbar onset were observed for any of the proteins (**Fig. 1C**).

**Figure 1:**
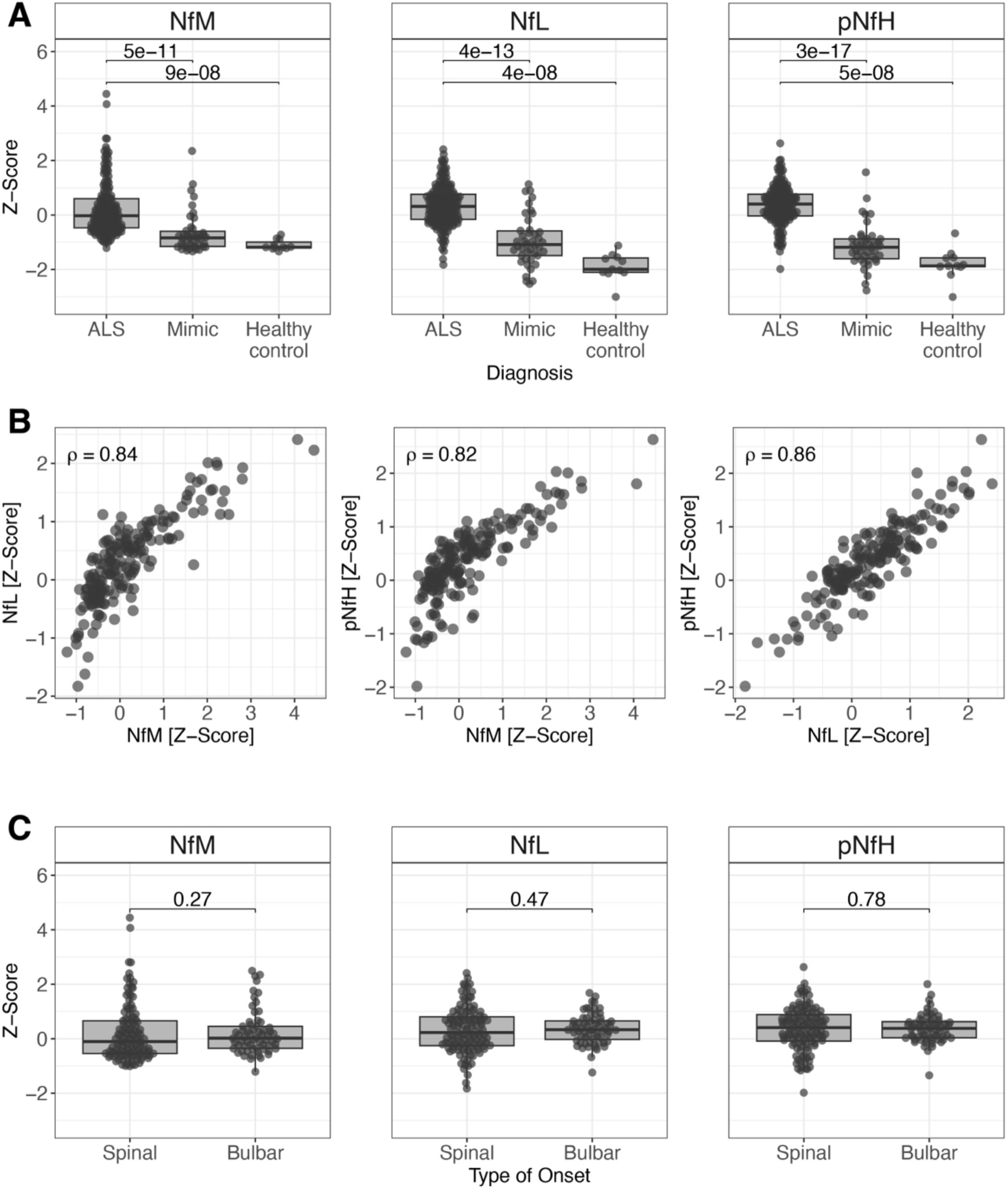
CSF levels of NfM, NfL and pNfH in relation to diagnosis and disease onset. (A) Levels of all three neurofilament proteins were significantly increased in patients with ALS compared with ALS mimics and healthy controls. (B) Strong correlations among patients with ALS were observed between all three proteins. (C) No difference in protein levels was found between patients with spinal and bulbar onset.

### High NfM levels are associated with fast disease progression

Comparing NfM levels between patients with different disease progression rates revealed significantly increased levels in fast progressors compared with slow progressors, reflecting the results for NfL and pNfH (NfM p=3e-06, NfL p=4e-06, pNfH p=4e-06) (**Fig. 2A**). This was also noted in the prediction of progression rate, which was evaluated using generalised linear models, ROC curves and AUC. While no statistical differences were found between the obtained ROC curves, we observed that a combination of all three proteins, as well as protein pairs such as NfM with pNfH, and NfL with pNfH, resulted in the highest AUC (AUC=0.87). Similar AUC values were observed for combinations of NfL with NfM and for NfM alone (AUC=0.86). The classification ability of NfL and pNfH alone were lower but also in the same range (AUC=0.85) (**Fig. 2B**).

**Figure 2:**
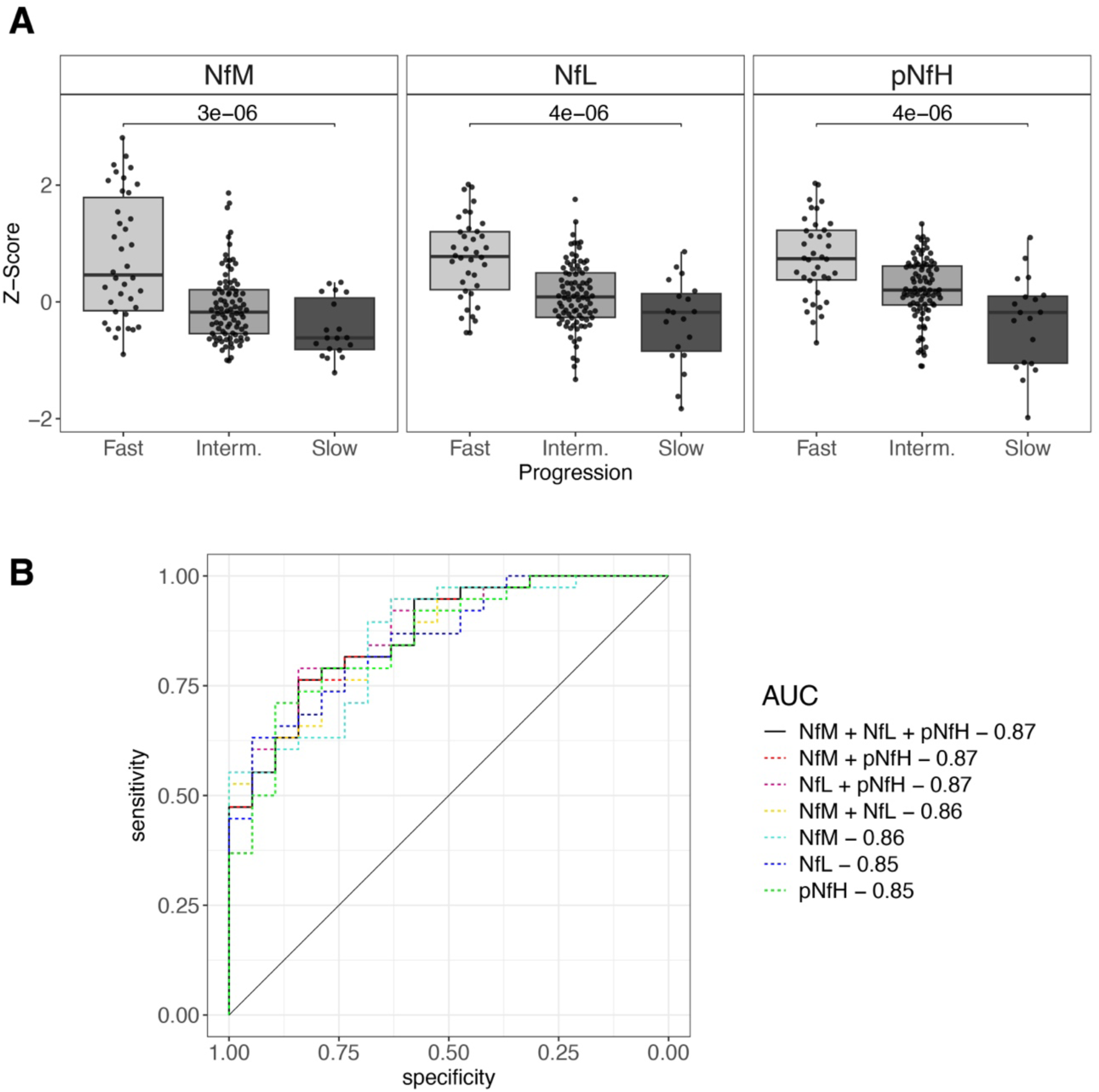
CSF neurofilament levels in patients stratified based on progression rate. (A) Significantly higher CSF levels were found in the group with fast progressors for all three proteins. (B) Receiver operating characteristic curve showing the neurofilaments ability to separate fast and slow progressors, separately or in combination, based on CSF protein levels.

Furthermore, we examined the correlation between protein levels and progression rate groups per ALSFRS-R subscores. We observed significant differences in NfM levels between fast and slow progressors across the subscores bulbar, fine motor, and gross motor. A similar result was also seen for pNfH, while for NfL, also the respiratory subscore displayed significant differences between fast and slow progressors (**Fig. 3A**). Additionally, the correlations to disease progression rates revealed moderate but statistically significant associations for all proteins and subscores (**Fig. 3B**), with *rho* ranging between 0.36-0.55 for bulbar, fine motor and gross motor, and between 0.18-0.21 for respiratory subscores.

**Figure 3:**
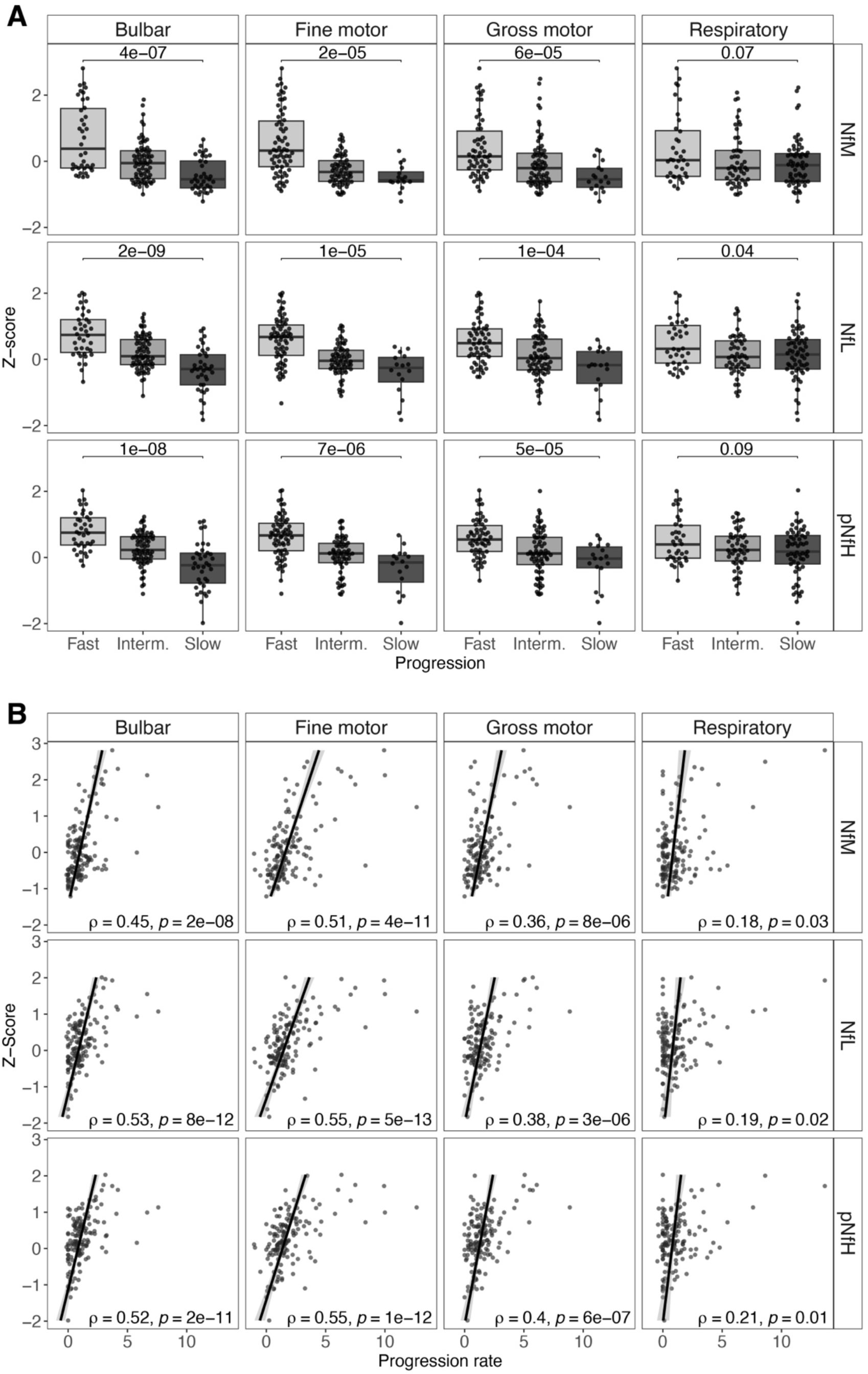
Protein levels in progression groups for the four ALSFRS-R subscores and correlations of protein levels with disease progression rate. (A) Significant differences between fast and slow progressors were observed for all proteins in bulbar, fine motor, and gross motor subscores. Only NfL showed significant differences between fast and slow progressors for the respiratory subscore. (B) Moderate but significant correlations were found between average disease progression and proteins levels for all proteins in all four subscores.

Next, we explored whether there was a difference in protein levels among individuals with a spinal onset compared with individuals with a bulbar onset, within the progression groups for the different ALSFRS-R subscores. While no differences were observed for NfM in any of the progression groups, significant differences were found between spinal and bulbar onset among fast progressors within the bulbar subscore for NfL (p=0.02) and pNfH (p=0.02), revealing increased levels in those with spinal onset. In the case of intermediate progressors, differences were identified within the bulbar subscore for pNfH (p=0.03), where higher levels were observed in individuals with spinal onset, and in the fine motor subscore for NfL (p=0.03) for which increased levels were observed in individuals with bulbar onset. Within the slow progressors, a significant difference was observed only within the fine motor subscores for NfL (p=0.02), with higher levels among the individuals with bulbar onset (**Supplementary Fig. 1**).

### High NfM levels are associated with shorter survival

The survival time for the patients with ALS included in this study ranged from 1 to 84 months. In the survival analysis, patients were divided into three groups based on protein levels, where the upper tercile (high) and lower tercile (low) were included in the following analysis. The Kaplan-Meier curves showed a clear separation between the groups, with higher protein levels associated with lower probability of survival for all three proteins (NfM p=1e-04; NfL p=9e-06; pNfH=3e-04) (**Fig. 4**, **Table 3**). The association between protein levels and survival probability was consistent regardless of onset site (spinal onset: NfM p=0.002; NfL p=9e-04; pNfH=0.004, bulbar onset: NfM=0.01; NfL=6e-04; pNfH=2e-04) (**Fig. 4**).

**Figure 4:**
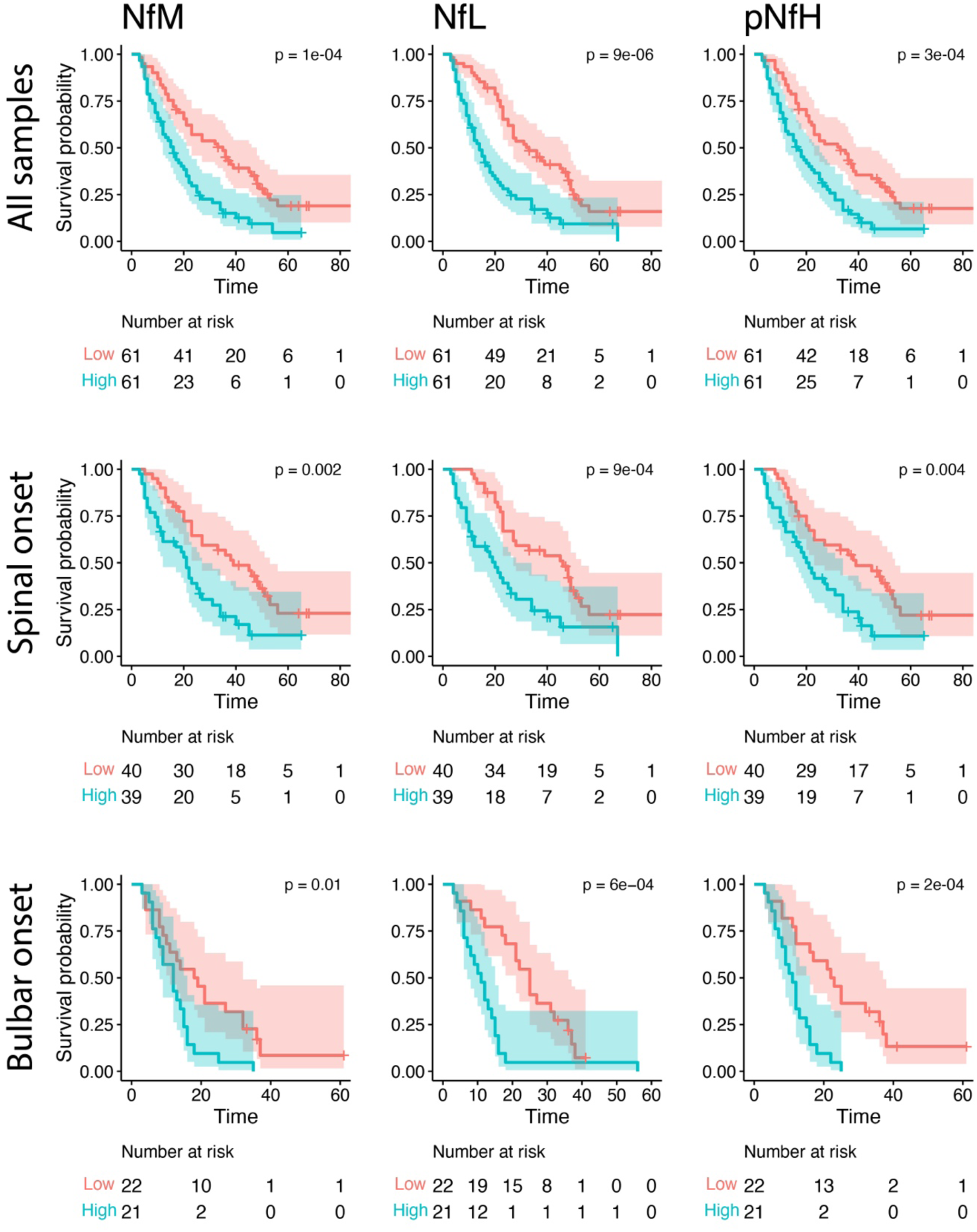
Kaplan-Meier survival curves illustrating survival probabilities in patients grouped based on protein levels. Lower levels of neurofilament proteins were associated with higher survival probability in all patients combined, as well as when bulbar and spinal onset patients were analysed separately.

**Table 3:** log rank p-values from survival analysis using Kaplan-Meier curves, comparing patients with ALS with high and low protein levels, in patients with spinal and bulbar onset separately. In all comparisons, a higher survival probability was observed in patients with low levels of the proteins.

| **Protein** | **Onset** | **p-value** |
| --- | --- | --- |
| **NfM** | *All* | 0.0001 |
|  | *Spinal* | 0.002 |
|  | *Bulbar* | 0.01 |
| **NfL** | *All* | 0.000009 |
|  | *Spinal* | 0.0009 |
|  | *Bulbar* | 0.0006 |
| **pNfH** | *All* | 0.0003 |
|  | *Spinal* | 0.004 |
|  | *Bulbar* | 0.0002 |

## Discussion

In this study, our objective was to explore the levels of NfM in CSF from 182 patients with ALS, 42 ALS mimics, and 11 healthy controls. Specifically, we aimed to evaluate the potential of NfM levels in reflecting ALS disease progression rates and survival probability, and compare the results to NfL and pNfH. For this, we included a subset of previously published NfL and pNfH data (11) and supplemented it with newly generated NfM data.

We observed increased levels of NfM in patients with ALS compared with mimics and controls, and a slight increase in mimics compared with healthy controls, much in line with previous reports on NfL and pNfH (4–9, 28). Results for the three subunits showed a strong correlation among patients with ALS, with coefficients ranging from ρ=0.82 (pNfH-NfM) to ρ=0.86 (NfL-pNfH), also aligning with prior findings (6, 29, 30). Although the correlations were strong, comparisons including NfM did not show a linear trend indicating that the linear ranges of the used assays were not identical.

When investigating the levels of NfM in relation to disease progression, we found higher levels in fast progressing patients compared with slow progressors. This trend was similar for NfL and pNfH, which concurs with previous studies (8, 12, 31, 32). Additionally, all three proteins showed similar performance in classifying fast progressors from slow progressors, and using two or three proteins in combination only improved the results marginally. To the best of our knowledge, a comprehensive investigation of protein levels concerning the four subscores within the ALSFRS-R scale has not yet been reported. In our analysis, we observed that higher protein levels were associated with faster subscore progression for three out of the four subscores, namely bulbar, fine motor, and gross motor subscore. For the respiratory subscore, only NfL reached statistical significance between fast and slow progressors. Here it is important to note that other factors, such as the use of non-invasive ventilation, or influence of medications such as inhalations may also impact the score. Such variables was not accounted for in the analysis.

In the present study, higher NfM levels also associated with reduced survival probabilities similar to previous reports for NfL and pNfH (6, 12, 13). Further stratification based on the onset site (spinal or bulbar) showed that increased levels of each neurofilament protein were linked to shorter survival, regardless of site of onset.

The levels of neurofilaments in CSF have been widely studied in relation to neurodegenerative disorders. Although the proteins are commonly found increased in disease populations as compared to healthy controls (19, 33–35), a multi-disease systemic review and meta-analysis on NfL noted that CSF levels in ALS and ALS/FTD patients ranked among the highest across various investigated diseases (36). The increased levels of neurofilaments observed in patients with ALS, in contrast to other diseases, are hypothesized to result from the higher concentration of axonal proteins in motor neurons relative to other neuronal subtypes (37). It has also previously been reported that the stoichiometry of the three neurofilament subunits differed in patients with ALS compared with healthy individuals. Zucchi *et al*. reported that the average value for NfL:NfM:NfH levels in plasma were 24:2.4:1.6 for patients with ALS, compared with 7:3:2 in a control group (17). Another study formulated the adaptive protein stoichiometry hypothesis which suggests that the abundance of Nf subunits is adjusted to limit energy consumption and prevent neuronal death, favouring less energy expensive NfL over pNfH and NfM (38).

Some limitations with this study include that the mimic group comprises diverse diagnoses such as multiple sclerosis, Parkinson’s disease, spinal stenosis, benign fasciculations, etc. (**Supplementary table 1**), and in all these cases the initial symptoms closely resembled those of ALS. Although the diversity within the mimic group poses a challenge, it is important to use cohorts that reflect the clinical reality to identify biomarkers capable of differentiate between these mimics and patients with ALS. Such differentiation is not only important for ensuring accurate care and early treatment, but also for strategically allocating medical resources where they can be most effective. Further, the non-linear correlation between the NfM levels and the two other neurofilaments, together with the lack of absolute quantification of the NfM levels, prevents us from delving deeper into the relationship between the different subunits. This limitation hampers the ability to fully explore their potential as biomarkers, including their combined ratio. Such analysis could give valuable insight into the stoichiometry between the proteins and their potential implications for ALS pathology. Furthermore, the utilization of selected cut-off levels for defining progression groups, as well as for high and low protein levels in the Kaplan-Meier analysis, introduces another limitation. The cohort-specific determination of the cut-offs could impact the interpretation of these findings and comparability with other studies. The absence of longitudinal data in this study limits our ability to explore how changes in neurofilament levels over time may correlate with disease progression. Ultimately, due to the challenges faced in collecting CSF, only a restricted number of CSF samples from healthy controls was available for analysis. Addressing these limitations in future research would contribute to a better understanding of the relationships among neurofilament subunits and their dynamic role in ALS.

In summary, the findings of this study demonstrate that NfM exhibits patterns and efficacy comparable to NfL and pNfH, successfully distinguishing between patients with ALS and mimics, as well as healthy controls. Notably, NfM also shows comparable performance in predicting disease progression and survival probability. While NfM alone may not provide additional diagnostic or prognostic value, future efforts of more detailed assessment regarding the stoichiometry of NfL, NfM and pNfH holds the potential to deepen our understanding of ALS pathology. Moreover, this collective approach might enhance the clinical utility of neurofilament markers, providing valuable insights for advancing future research in ALS.

## Acknowledgements

We gratefully acknowledge the participants and thank them for their time and cooperation in this study.

## Data availability statement

Anonymized data can be obtained through reasonable requests to the corresponding author. Please note that the data is not publicly available, as it is subject to privacy and ethical restrictions.

