## Supplemental material for "Cerebrospinal fluid levels of NfM in relation to NfL and pNfH as prognostic markers in amyotrophic lateral sclerosis"

***Correspondence to:**

Dr. Anna Månberg

Dept of Protein Science

ORCID: 0000-0002-0056-1313

### Supplementary Tables

Table 1: List of diagnoses of individuals in the ALS mimic group.

| Diagnosis |
|---|
| Adult-onset leukoencephalopathy with axonal spheroids and pigmented glia |
| Benign fasciculations |
| Bulbar dystonia |
| Cervical radiculopathy |
| Degenerative spine |
| Dysarthria and dysphagia |
| Familial prion disease |
| Inclusion body myositis |
| Isolated unilateral hypoglossal nerve paralysis |
| Lumbar spondylosis |
| Motor neuropathy |
| Multiple sclerosis |
| Multiple system atrophy (parkinsonian type) |
| Myelopathy |
| Neuroborreliosis |
| Paraneoplastic process |
| Parkinson's disease |
| Polymyositis |
| Polyneuropathy |
| Progressive multifocal leukoencephalopathy |
| Progressive supranuclear palsy |
| Psychological stress and fatigue |
| Spastic paraparesis |
| Spinal stenosis |
| Suspected mitochondrial disease |
| Vasculitic neuropathy |

### Supplementary Figures

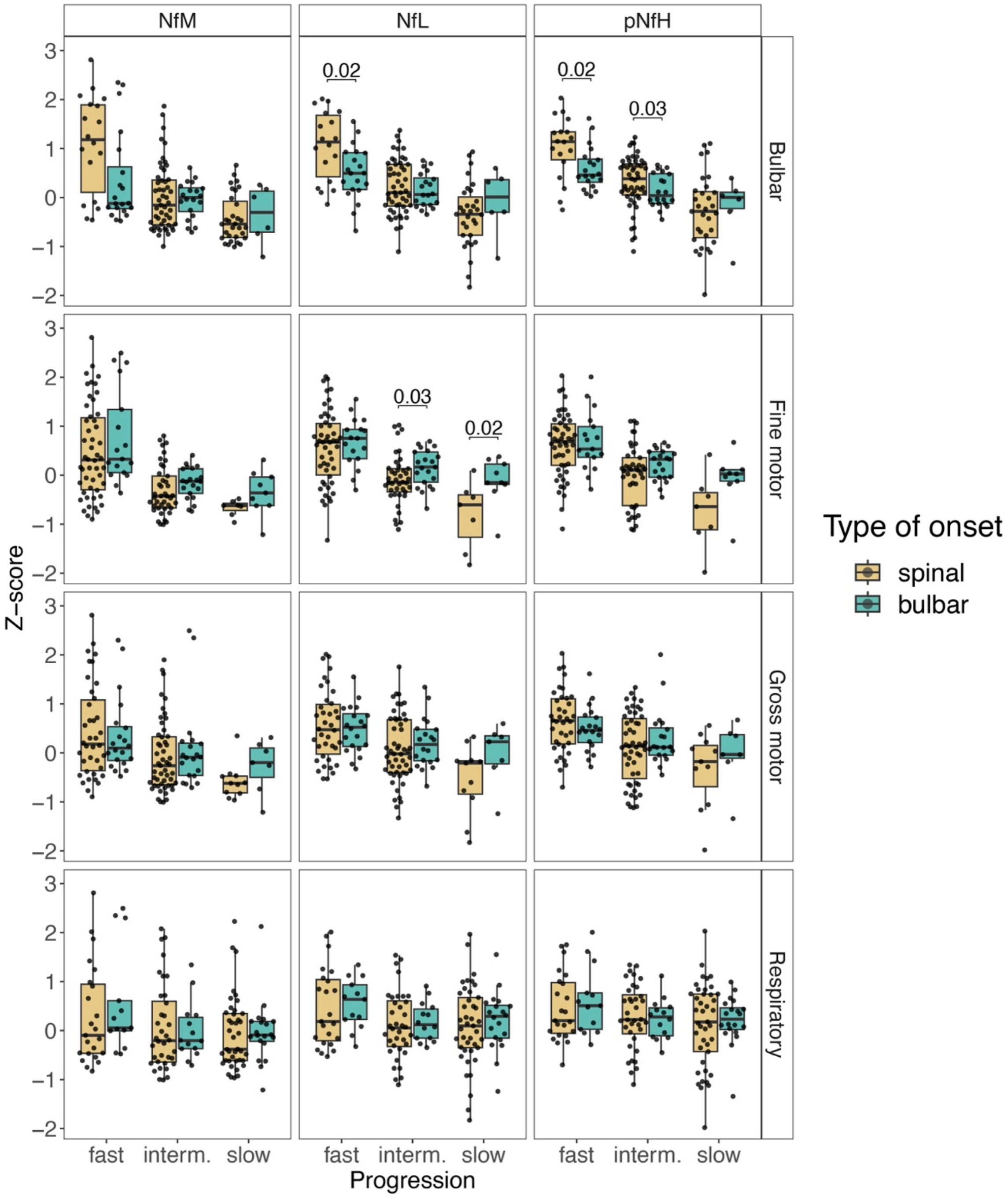


Supplementary Figure 1: Differences in CSF protein levels between patients with spinal and bulbar onset within each progression group for each protein and ALSFRS-R subscore.
